# Worsening pre-existing health conditions in U.S. adults with Long COVID

**DOI:** 10.64898/2026.09.22.26363331

**Authors:** Deja L Edwards, Alexandra F Rollins, Nicole D Ford, Caroline Pratt, Robin Bloodworth, Kristen Pogreba-Brown, Resa M. Jones, Vinita Sharma, Miguel Reina Ortiz, Sharon Saydah

**Affiliations:** Eagle Global Scientific, Huntsville, AL, 35806, USA; Coronavirus & Other Respiratory Viruses Division (CORVD), National Center for Immunization and Respiratory Diseases (NCIRD), Centers for Disease Control and Prevention (CDC), 1600 Clifton Rd., NE, Mailstop US1-1, Atlanta, GA 30329-4027, USA; Abt Global Inc., 6130 Executive Blvd, Rockville, MD 20852; Department of Epidemiology and Biostatistics, Zuckerman College of Public Health, University of Arizona, Tucson, AZ; Department of Epidemiology and Biostatistics, College of Public Health, Temple University, Philadelphia, PA; Fox Chase Cancer Center, Temple Health, Temple University, Philadelphia, PA; Department of Community and Global Health, Richard M. Fairbanks School of Public Health, Indiana University, Indianapolis, IN, 46202, USA

## Abstract

**Purpose:** Long COVID literature often describes new-onset symptoms or conditions following SARS-CoV-2 infection; however, Long COVID also encompasses the exacerbation of pre-existing conditions. We describe health conditions reportedly worsened by COVID-19 illness and associated concerns in U.S. adults.

**Methods:** We collected longitudinal data from adults 18 years and older participating in the Track Post COVID-19 Conditions, an active Long COVID cohort surveillance project. Respondents completed surveys 3-, 6-, and 12- months after COVID-19 illness, beginning in July 2023. In each survey, respondents were asked whether they had any health conditions or concerns that were made worse by COVID-19. Those with an affirmative response were asked to specify the worsening condition(s) via free text. Some respondents used this free text to describe challenges or experiences related to their worsening conditions. This report categorizes and quantifies reported conditions and concerns by body system categories and qualitatively describes associated challenges and experiences using direct quotes from respondents’ free text responses.

**Results:** Among 2,227 adults, 263 (11.8%) respondents reported at least one worsening condition or concern following COVID-19 illness. Of these, nearly 32% reported a worsening respiratory condition or concern. Adults with worsening pre-existing conditions reported challenges accessing adequate care, completing daily activities, and managing their symptoms following COVID-19.

**Discussion:** While Long COVID literature often focuses on the development of new-onset long-term symptoms, more than 1 in 10 adults reported worsening of a pre-existing condition following COVID-19. Reported challenges with accessing care and daily activities highlight potential need for accommodations or support for adults with Long COVID.

## Introduction

Certain chronic and pre-existing health conditions increase the risk a patient will develop Long COVID and other poor health outcomes following acute COVID-19 [1–3]. Long COVID literature often describes the range of new-onset symptoms presenting or persisting for months following SARS-CoV-2 infection, but Long COVID also encompasses the exacerbation of pre-existing conditions [4–5]. Further, studies have shown that Long COVID is more common among people with pre-existing conditions [5–6]. A study comparing data from the 2022 Household Pulse Survey and the 2022 National Survey on Health and Disability found the prevalence of Long COVID was significantly higher among people with pre-existing health conditions and disabilities than the general U.S. population [6]. Individuals living with Long COVID often describe experiencing multiple symptoms simultaneously [7] with subsequent impact on their social and professional lives [6–9]. Experiences and challenges navigating the healthcare system have also been shared by patients with Long COVID [8; 10]. Less is known in the U.S. about the challenges and burden of Long COVID experienced by those whose pre-existing conditions worsen after COVID-19.

Studies researching the long-term impact of COVID-19 on pre-existing conditions often focus on one or a small set of health conditions and are not comprehensive in the conditions included within the analysis [11–16]. For example, after SARS-CoV-2 infection, patients with pre-existing neurological conditions were more likely to develop an altered mental status, fatigue, and other poor outcomes [11], and those with Alzheimer’s disease and Parkinson’s disease experienced worsened overall outcomes, including exacerbation of motor and cognitive symptoms and accelerated disease progression [12]. Individuals with pre-existing cardiovascular conditions [13–14], diabetes [15], and certain respiratory conditions [16] may also experience worsening of their condition(s) following COVID-19. However, these previous studies are limited to specific health conditions, are based on cross-sectional studies, or include only patients who were hospitalized during their acute COVID-19 illness.

To better understand exacerbation of chronic conditions following COVID-19, we analyzed prospectively collected, longitudinal survey data from U.S. adults describing health concerns and conditions reportedly exacerbated by COVID-19 and participant experiences with worsening conditions.

## Methodology

### Study design and sample

The Tracking the Burden, Distribution, and Impact of Post COVID-19 Conditions (Track PCC) active surveillance project examines the burden and long-term health impacts following COVID-19 illness by collecting longitudinal survey data from adults 3, 6, and 12 months following a SARS-CoV-2 infection. Details about the Track PCC study design and methods can be found elsewhere [17]. Briefly, active surveillance data come from four sites – Arizona, Indiana, the Bronx (New York), and North/Northeast Philadelphia (Pennsylvania). Respondents were recruited to participate in the 3-month (i.e., baseline) survey between 70-112 days after they had a COVID-19 illness (diagnosed via home-test kit; lab test/PCR; or test at a clinic, urgent care, or a doctor’s office). This analysis was limited to adults aged 18 years or older at the time of the COVID-19 illness. Response rates, where available, varied by site from 19-86%. Data for this analysis were collected during July 9, 2023–January 15, 2025. Respondents provided informed written consent prior to participating. More details on the survey design and questions are described below.

Surveys were completed primarily online using REDCap; one site also allowed respondents to complete their surveys via U.S. mail or by telephone. Participation was voluntary, and respondents were able to skip questions or discontinue participation at any time.

For these analyses, respondents were included if they 1) completed, at a minimum, the Track PCC 3-month survey; and 2) reported any worsening condition or concern after COVID-19 in one or more surveys, for a total of 263 of 2,227 adults enrolled in Track PCC (Supplemental Materials). Respondents may have reported multiple worsening conditions on each survey; data from all surveys (3-, 6-, and 12-month) were included in analysis.

### Survey Questions

#### Demographics and pre-existing health conditions

On the 3-month survey, participants reported their date of birth, sex (male or female), and race (Asian or Asian American, Black or African American, Native Hawaiian or other Pacific Islander, American Indian or Alaska Native, White or Caucasian, any Other race – please specify) and ethnicity (Hispanic or non-Hispanic) from the options provided. Respondents were also asked whether they had ever been told by a doctor or health professional they had any of the following pre-existing conditions before the index date of their COVID-19 illness: chronic lung disease, mental health disorder, immune system disorder, cancer, chronic kidney disease, chronic liver disease, chronic headaches, heart disease, high blood pressure, high cholesterol, diabetes, gestational diabetes, chronic pain, stroke, or obesity. Respondents were also able to self-report any other pre-existing health conditions using a free text response.

#### Worsening conditions or concerns after COVID-19

On each survey (i.e., 3-, 6-, and 12-months following COVID-19 illness), respondents were asked, “do you have any health conditions or concerns that have been made worse by COVID-19?” (yes/no). These data were collected separately from the pre-existing health conditions described above, meaning the conditions and concerns reported as worsening following COVID-19 may or may not have been reported as a clinically diagnosed pre-existing condition. Those who responded affirmatively were then asked to use free text response to describe those health conditions or concerns. Some respondents used this free text to describe challenges and the impacts these worsening conditions may have had on their lives. Respondents were able to report more than one worsening condition.

### Analysis

Both quantitative and qualitative methods were used in this analysis. We used descriptive statistics to summarize demographic characteristics of survey respondents who reported any worsening condition(s) or concern(s). Additional information about the demographics of the overall Track PCC cohort participants is provided in the Supplemental Materials (Supplemental Table 1).

Two researchers independently reviewed each free-text response describing health conditions or concerns that were made worse by COVID-19 and compiled a list. Respondents may have used clinical diagnoses or lay terms (e.g., brain bleed, heart racing) to describe their conditions in the free text; we retained the language they used. A clinician reviewed the complete list of reported worsening conditions or concerns and categorized them into body system categories and, when appropriate, subcategories. The body system categories these conditions and concerns were assigned to were as follows: *cardiovascular/cerebrovascular, dermatologic, endocrine/metabolic, GI (gastrointestinal), EENT (ears, eyes, nose, and throat), immune/autoimmune, mental health, musculoskeletal, neurologic, neuropsychiatric, respiratory, general (e.g., fatigue), urinary, and other health conditions*. Subcategories were utilized to precisely identify sets of conditions and concerns reported by respondents, when appropriate. The complete list of conditions or concerns grouped by body system and subcategory are described in the Appendix.

We present the number and proportion of respondents reporting any worsening condition or concern by body system category (cardiovascular/cerebrovascular, dermatologic, etc.). The number of responses in each subcategory are also presented. Because respondents were able to report more than one worsening condition or concern, the sum of responses for subcategories may exceed the number reported for the overall conditions by body system.

We further reviewed the free text responses to inductively identify common themes. Themes within the free-text responses often touched on one of four key areas often discussed in Long COVID literature: impact on daily activities, management of worsening conditions and concerns, seeking medical care for concerns, and experiencing multiple worsening concerns [11; 18]. Based on these themes, we reviewed each response and selected representative quotes related to each theme as narrative examples. We included an example from each body system category representing 10% or more of the sample.

This activity was reviewed by CDC, deemed not research, and was conducted consistent with applicable federal law and CDC policy (see e.g., 45 C.F.R. part 46.102(l)(2), 21 C.F.R. part 56; 42 U.S.C. §241(d); 5 U.S.C. §552a; 44 U.S.C. §3501 et seq.). Prior to project implementation, the protocol, appendices, and other associated documents were reviewed and approved by the IRB overseeing each surveillance site’s project activities in accordance with their own policies and procedures. An exemption from review was obtained by all sites.

## Results

### Respondent characteristics

Sociodemographic characteristics of the analytic sample are presented in Table 1. Over half (55.4%) of adult respondents were white, 77.8% were female, the median age was 57 [IQR 43-67] years, and 93.5% of respondents reported a pre-existing condition before their COVID-19 illness. The characteristics of the worsening conditions analytic sample were similar to those of the overall Track PCC cohort (Supplemental Table 1).

**Table 1.** Demographic characteristics of adult respondents who reported worsening of any pre-existing condition or concern after COVID-19.

|  | **N** |
| --- | --- |
| **Adults reporting any worsening health conditions or concerns<sup>1</sup>** | 263 |
| **Age, years (IQR)** | 57 (43, 67) |
| **Age group** |  |
| 18-49 years | 89 (33.8) |
| 50-64 years | 96 (36.5) |
| 65+ years | 78 (29.7) |
| **Site** |  |
| Arizona | 44 (16.7) |
| The Bronx, New York | 16 (6.1) |
| Indiana | 70 (26.6) |
| North/Northeast Philadelphia | 133 (50.6) |
| **Sex** |  |
| Male | 58 (22.2) |
| Female | 203 (77.8) |
| Missing | 2 |
| **Race** |  |
| Black or African American | 67 (26.7) |
| White or Caucasian | 139 (55.4) |
| Any Other Race or Multiple Races | 45 (17.9) |
| Missing | 13 |
| **Ethnicity** |  |
| Hispanic | 55 (22.4) |
| Non-Hispanic | 191 (77.6) |
| Missing | 17 |
| **Pre-existing condition<sup>2</sup>** |  |
| Any underlying condition | 246 (93.5) |

<sup>1</sup>Responses are collected from 3- (baseline), 6-, and 12-month surveys. Respondents may have reported multiple conditions or concerns on each survey.
<sup>2</sup>All respondents were asked whether they had any worsening health conditions or concerns, regardless of whether they reported any pre-existing health conditions. As a result, some of the worsening health conditions or concerns reported may not have been diagnosed by a healthcare provider and therefore were not reported as a pre-existing condition.

### Worsening health conditions and concerns following COVID-19

Worsening health conditions and concerns following COVID-19 are presented in Table 2. Worsening conditions in the respiratory (31.9%), general (20.2%), and musculoskeletal (14.8%) body system categories were the most commonly reported (Table 2). Among the sub-categories of worsening conditions or concerns, lung diseases and conditions (e.g., asthma and COPD) (n=42), respiratory signs/symptoms (e.g., cough and shortness of breath) (n=40), body aches/pain (n=37), and anxiety and depression (n=27) were the most frequently reported. Some conditions or concerns were less often reported, like those in the endocrine (n=7) body system category, which included diabetes (n=3) and thyroid concerns (e.g., hypothyroidism and Hashimoto’s) (n=3) subcategories. Several conditions or concerns, such as sarcoidosis, endometriosis, and hives, were only reported by a single respondent.

**Table 2.**
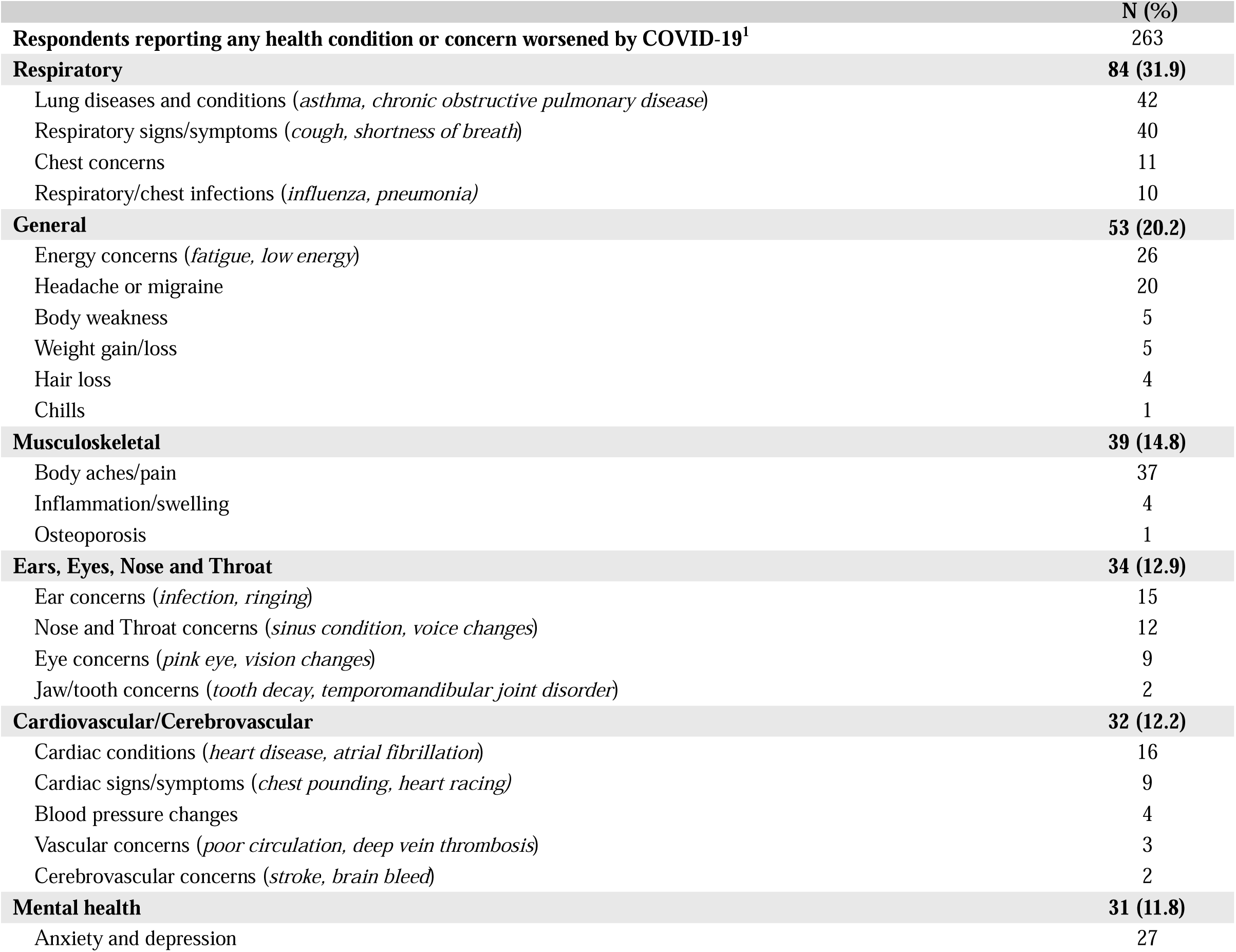
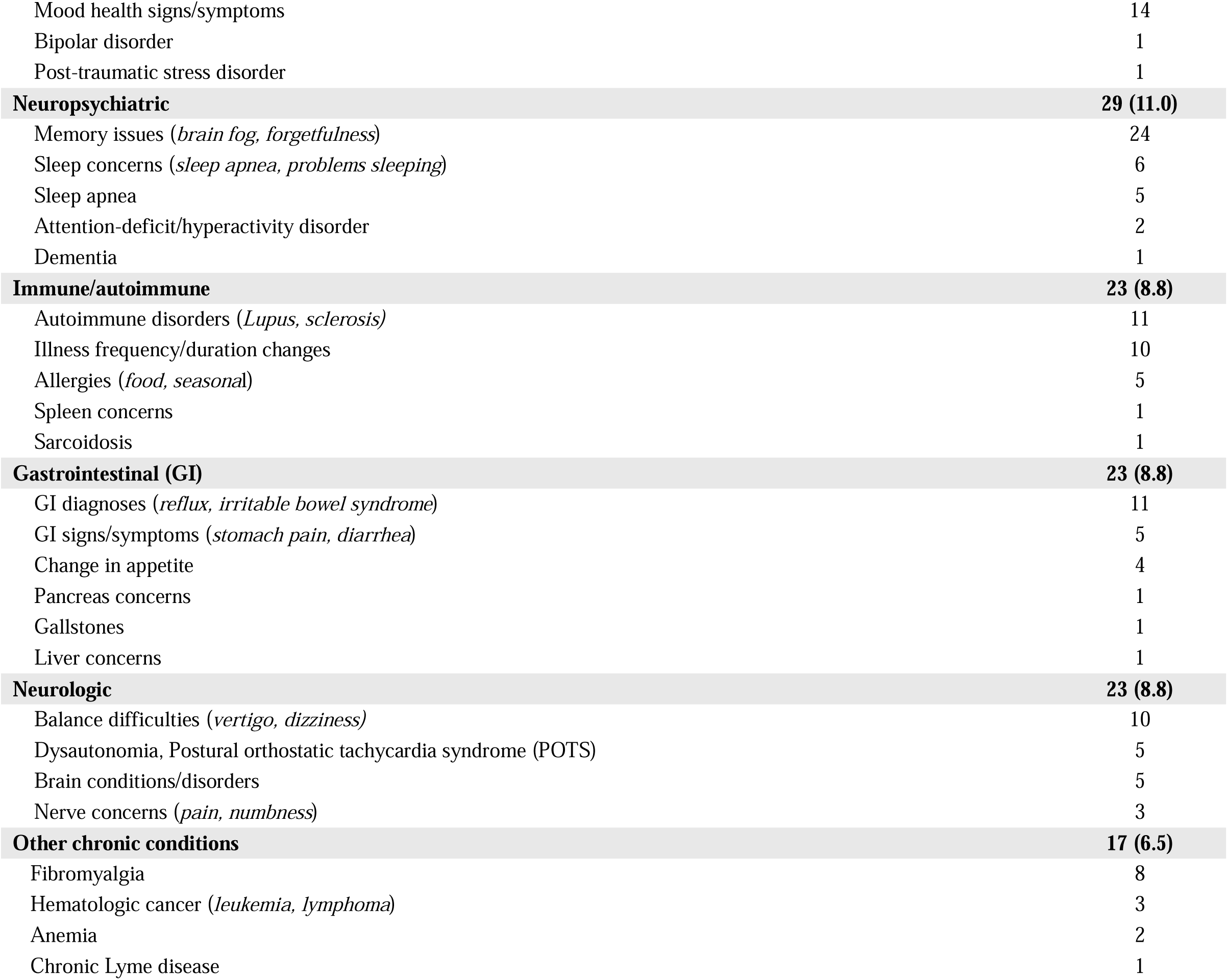
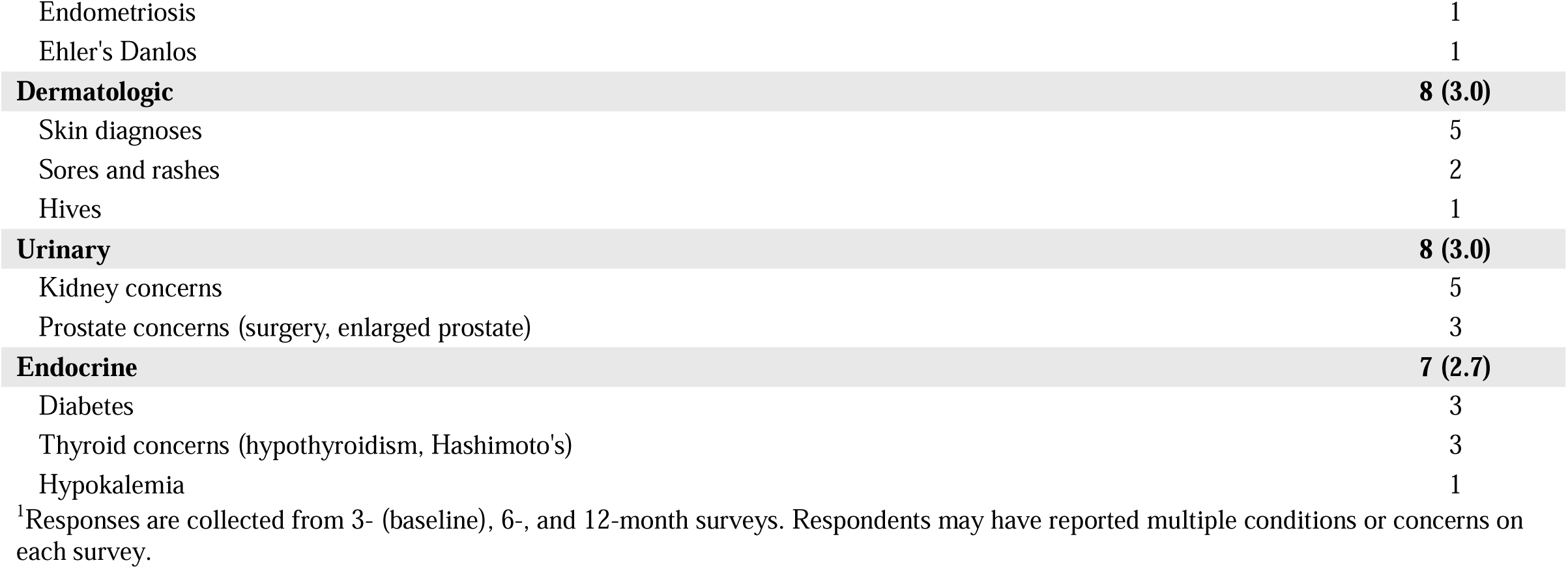
Worsening health conditions and concerns after COVID-19 reported by adult respondents via free text response.

|  | N (%) |
| --- | --- |
| **Respondents reporting any health condition or concern worsened by COVID-19<sup>1</sup>** | **263** |
| **Respiratory** | **84 (31.9)** |
| Lung diseases and conditions (*asthma, chronic obstructive pulmonary disease*) | 42 |
| Respiratory signs/symptoms (*cough, shortness of breath*) | 40 |
| Chest concerns | 11 |
| Respiratory/chest infections (*influenza, pneumonia*) | 10 |
| **General** | **53 (20.2)** |
| Energy concerns (*fatigue, low energy*) | 26 |
| Headache or migraine | 20 |
| Body weakness | 5 |
| Weight gain/loss | 5 |
| Hair loss | 4 |
| Chills | 1 |
| **Musculoskeletal** | **39 (14.8)** |
| Body aches/pain | 37 |
| Inflammation/swelling | 4 |
| Osteoporosis | 1 |
| **Ears, Eyes, Nose and Throat** | **34 (12.9)** |
| Ear concerns (*infection, ringing*) | 15 |
| Nose and Throat concerns (*sinus condition, voice changes*) | 12 |
| Eye concerns (*pink eye, vision changes*) | 9 |
| Jaw/tooth concerns (*tooth decay, temporomandibular joint disorder*) | 2 |
| **Cardiovascular/Cerebrovascular** | **32 (12.2)** |
| Cardiac conditions (*heart disease, atrial fibrillation*) | 16 |
| Cardiac signs/symptoms (*chest pounding, heart racing*) | 9 |
| Blood pressure changes | 4 |
| Vascular concerns (*poor circulation, deep vein thrombosis*) | 3 |
| Cerebrovascular concerns (*stroke, brain bleed*) | 2 |
| **Mental health** | **31 (11.8)** |
| Anxiety and depression | 27 |
| Mood health signs/symptoms | 14 |
| Bipolar disorder | 1 |
| Post-traumatic stress disorder | 1 |
| **Neuropsychiatric** | **29 (11.0)** |
| Memory issues (*brain fog, forgetfulness*) | 24 |
| Sleep concerns (*sleep apnea, problems sleeping*) | 6 |
| Sleep apnea | 5 |
| Attention-deficit/hyperactivity disorder | 2 |
| Dementia | 1 |
| **Immune/autoimmune** | **23 (8.8)** |
| Autoimmune disorders (*Lupus, sclerosis*) | 11 |
| Illness frequency/duration changes | 10 |
| Allergies (*food, seasonal*) | 5 |
| Spleen concerns | 1 |
| Sarcoidosis | 1 |
| **Gastrointestinal (GI)** | **23 (8.8)** |
| GI diagnoses (*reflux, irritable bowel syndrome*) | 11 |
| GI signs/symptoms (*stomach pain, diarrhea*) | 5 |
| Change in appetite | 4 |
| Pancreas concerns | 1 |
| Gallstones | 1 |
| Liver concerns | 1 |
| **Neurologic** | **23 (8.8)** |
| Balance difficulties (*vertigo, dizziness*) | 10 |
| Dysautonomia, Postural orthostatic tachycardia syndrome (POTS) | 5 |
| Brain conditions/disorders | 5 |
| Nerve concerns (*pain, numbness*) | 3 |
| **Other chronic conditions** | **17 (6.5)** |
| Fibromyalgia | 8 |
| Hematologic cancer (*leukemia, lymphoma*) | 3 |
| Anemia | 2 |
| Chronic Lyme disease | 1 |
| Endometriosis | 1 |
| Ehler's Danlos | 1 |
| **Dermatologic** | **8 (3.0)** |
| Skin diagnoses | 5 |
| Sores and rashes | 2 |
| Hives | 1 |
| **Urinary** | **8 (3.0)** |
| Kidney concerns | 5 |
| Prostate concerns (surgery, enlarged prostate) | 3 |
| **Endocrine** | **7 (2.7)** |
| Diabetes | 3 |
| Thyroid concerns (hypothyroidism, Hashimoto's) | 3 |
| Hypokalemia | 1 |

<sup>1</sup>Responses are collected from 3- (baseline), 6-, and 12-month surveys. Respondents may have reported multiple conditions or concerns on each survey.

### Qualitative description of worsening conditions or concerns

Sample quotes from respondents describing how their worsening condition(s) following COVID-19 illness has affected their lives are presented in Table 3. They often described limitations living with these conditions or concerns and how they “have been [worse] than before [their] first diagnosis of COVID.” Limitations varied by reported condition or concern; for example, respondents with worsening musculoskeletal concerns including joint and muscle pains often reported challenges with their physical abilities like walking and balancing. Respondents shared sentiments of daily limitations “do[ing] the social and physical activities that [they] once loved…often feel[ing] discouraged to do things because of” their worsening condition or concern. Fears of re-infection and further exacerbations of their condition or concern also impacted respondents’ mental wellbeing. “Sometimes I choose not to do work things because of fear of infection. [Visual vertigo] really impacted my ability to do my job this year. With each re-infection, I am so scared that I will regress really bad or develop new long symptoms.”

**Table 3.** Sample quotes from adult respondents describing their worsening conditions or concerns after COVID-19.

|  | **General description of worsening condition or concern** | **Impact on daily activities** | **Management of worsening conditions** | **Seeking medical care for concerns** | **Experience with multiple worsening concerns** |
| --- | --- | --- | --- | --- | --- |
| **Respiratory** | "Interstitial lung disease-coughing got worse" (F, 80's) | "I now have trouble breathing while doing activities or exercise." (F, 40's) | "Since COVID I have not been able to control my asthma" (F, 40's) | "I get pneumonia every year...I was in ICU for a month for respiratory failure, heart failure aspiration, seizures. I was on respirator, ventilator and the tracheostomy." (F, 30's) | "Chronic fatigue, sore throats and it seems like my asthma isn't controlled anymore" (F, 30's) |
| **General** | "I already had dysautonomia and chronic fatigue syndrome. Now everything is exacerbated and the gains I had made over the years are gone." (F, 60's) | "I have less energy to do the social or physical activities that I once loved. and often feel discouraged to do things because of my dizziness and nausea." (F, 30's) | "problems with migraines that cause dizziness and I am still dealing with those and trying to keep them under control." (F, 30's) | "Chills - I lost weight. I got real sick. Had to stay in the hospital." (F, 60's) | "pre-existing fatigue, poor memory/concentration, and IBS seem worse than before this last bout of COVID" (F, 70's) |
| **Musculoskeletal** | "Back pain has greatly escalated since COVID." (F, 40's) | "Problems with bottom of my feet - the best way to describe it is swelling on the balls of my feet. [My] left foot [is] leaning inward causing pain in ankle, especially when wearing shoe. Difficulty with balance." (F, 70's) | "Body pain - legs and feet lock up on me. Arms and fingers tingling/numb. I have to shake my legs to walk around" (F, 50's) | "blood clots in kidneys caused extreme back pain. Given blood thinners at hospital." (F, 20's) | "Body aches and headaches mainly. It felt like this started to become more regular since my positive test; usually I just get a cold but now I get a cold with COVID-like symptoms and am out for 2-3 days but test negative [for COVID]." (F, 20's) |
| **Eyes, Ears, Nose, Throat (EENT)** | "I already had severe sinus condition prior to COVID-19, COVID-19 made it worse" (F, 60's) | "My voice, I can't sing like I used to." (F, 70's) | "Since having COVID I am more prone to ear infections and the length of time it takes for my ears to clear of congestion after a cold. It took almost 2 months for my ears to completely drain and the "clicking" to stop after the persistent ear infections after COVID." (F, 60's) | "Sometimes I choose not to do work things because of fear of infection, and repeated re-infections really impacted my ability to do my job this year because symptoms last a long time for me. I got Post COVID permanent visual vertigo after my first infection. It disabled me for months until I could get PT and slowly start to heal but it will never be gone. I have been susceptible to getting COVID. With each reinfection I am so scared that I will regress really bad or develop new long symptoms. (F, 40's) | "I have TMJ and a tube in left ear. Have had an infection there since COVID" (M, 50's) |
| **Cardiovascular/**<br>**Cerebrovascular** | "Increased heart palpitations" (F, 50's) | * | "Very erratic blood pressures - took months to control" (F, 70's) | "I have atrial fibrillation and now I'm on heart medication, blood thinners and I have a heart monitor implanted" (F, 50's) | "My heart palpitations and blood pressure dropping (causing fainting) have become much more severe since I had COVID." (F, 60's) |
| **Mental health** | "My mental health has been off so much more than before my first diagnosis of COVID" (F, 30's) | "Seems like since I had COVID I've never been the same. I don't have the drive I had for me yard work. For me it [was] therapeutic. Now it's a chore... almost everything is a chore" (M, 70's) | <sup>1</sup> | <sup>1</sup> | "I have been experiencing the worse anxiety I have ever had to point where I'm having constant panic attacks. I'm having chest pain almost every single day" (F, 30's) |
| **Neuropsychiatric** | I have sleep apnea and insomnia and COVID-19 has made it 10x worse (F, 30's) | I have a very hard time thinking this out quickly. Processing normal conversations are a struggle for me since COVID, remembering directions is difficult as well as remembering where I am at sometime. (M, 60's) | <sup>1</sup> | <sup>1</sup> | ADHD symptoms have worsen[ed] especially short term memory, forgetfulness, emotional regulation (F, 20's) |

<sup>1</sup>Cells were left empty since none of the reviewed free text responses described both the condition category and limitation.

These challenges to respondents’ physical, social, and mental wellbeing were further complicated by experiencing multiple worsening conditions or concerns simultaneously. One respondent with a pre-existing neuropsychiatric condition stated their “ADHD symptoms have worsen[ed], especially short-term memory, forgetfulness, and emotional regulation.” In some instances, the worsening conditions or concerns affected multiple body systems. For example, one respondent had “pre-existing fatigue, poor memory/concentration, and IBS [that] seem[ed] worse than before [their] last bout of COVID.” Respondents reported utilizing medical interventions – visits to healthcare providers, hospital stays, medications, and COVID-19 vaccinations – to provide relief and support for their worsening conditions or concerns. Some utilized multiple forms of medical interventions, as described by one patient with atrial fibrillation who was “on heart medication, blood thinners, and [had] a heart monitor implanted.” Some respondents shared concerns that their condition or concern would never resolve or return to the state it was prior to their COVID-19 illness.

## Discussion

Long COVID may present as a new condition or an exacerbation of pre-existing conditions [19], yet most research has focused on incident conditions or symptoms. In the Track PCC active surveillance cohort, adults with various pre-existing conditions reported experiencing worsening of these conditions or concerns in the months following acute COVID-19 illness. Worsening conditions in the respiratory, general, and musculoskeletal body system categories were the most commonly reported. Lung diseases and conditions, respiratory signs/symptoms, body aches/pain, and anxiety and depression were the most commonly reported body system subcategories. Participants reported that these worsening conditions presented many challenges, including difficulties managing symptoms, having to address multiple worsening symptoms or conditions at once, and limitations in daily activities. It is important to gather qualitative information on worsening conditions to better understand patients’ experiences and barriers to better health. The intersection between pre-existing conditions and Long COVID highlights distinct challenges with respect to symptom management, healthcare navigation, and participation in social and work settings. Our findings may suggest that patients living with chronic conditions may benefit from follow-up with a healthcare provider to evaluate any exacerbations of their conditions and/or to provide additional support. Patient-centered care is likely important for comprehensive and long-term care of individuals with worsened pre-existing conditions following acute COVID-19 illness.

Participants in our study reported a wide range of worsening conditions and concerns — such as asthma, shortness of breath, body aches/pain, diabetes, and anxiety and depression. Although it is not well understood, some studies have reported that SARS-CoV-2 infection contributes to worsening neurological conditions, diabetes, and other pre-existing health conditions [11–15]. A study among patients from a clinic in Toledo, Ohio with postural orthostatic tachycardia syndrome (POTS) reported 68% experienced worsening of their symptoms in the months after their SARS-CoV-2 infection, most of whom required additional therapy to manage their worsening symptoms [30]. Similarly, 32% of patients with metabolic syndrome reported worsening hypertension requiring therapy in the 12 months following COVID-19 illness [31]. Patients with pre-existing irritable bowel syndrome (IBS) and other digestive concerns have also reported exacerbations of their symptoms, including constipation and concerns with their stool [32].

Worsening respiratory concerns and conditions were the most prevalent body system category reported by respondents in our study. Extensive literature has shown respiratory viruses contribute to exacerbations of pre-existing respiratory conditions, including chronic obstructive pulmonary disease (COPD) and asthma [20–24]. Although fewer studies have addressed the potential impacts of COVID-19 on worsening pre-existing respiratory conditions, evidence suggests COVID-19 may also exacerbate chronic respiratory conditions [25–26]. Data from the United Kingdom found nearly 80% of adults with pre-existing asthma experienced exacerbations in their difficulty breathing [27]. Evidence on the burden of exacerbation of chronic conditions after COVID-19 is not entirely conclusive and is lacking for most conditions [28–29].

Respondents in our study shared struggles with stress and social isolation as they live with chronic conditions and the risk of re-infection or newly worsening conditions. Some research suggests individuals with pre-existing mental health disorders may be at increased risk for worsening symptoms after a COVID-19 illness, although the data are not conclusive [33]. Social isolation, access to healthcare and resources, and availability of social support networks were mentioned as contributing factors that may impact how worsening mental health symptoms developed, which often varied by research study and may contribute to the inconclusive nature of the findings. Workers with chronic conditions returning to work during the pandemic shared similar concerns, fearing their risk of SARS-CoV-2 infection and potential barriers to their medical interventions leading to increased stress [34]. Some respondents in our study reported experiencing mental health concerns alongside other chronic conditions, suggesting worsening mental health concerns may be experienced simultaneously with other worsening health conditions, including fatigue, vision concerns, and other physical health concerns.

Long COVID literature rarely captures narratives of individuals living with or experiencing worsening conditions after COVID-19. In existing studies, adults often describe complications managing their condition, experiencing overlaps in symptoms, and needing medical intervention [19; 20; 27; 31]. Participants in Sullivan et al.’s study describe how “Long COVID has made a number of issues [they] already dealt with much worse” and their challenges as requiring “significant regular treatment (e.g., twice weekly physical therapy) to get closer to the baseline [they’d] established and been able to maintain fairly consistently for several years prior to contracting COVID” [19]. These stories from the people living with these complications may help highlight gaps in medical care, accessibility, and knowledge on Long COVID and more specifically how COVID-19 worsens pre-existing conditions.

Patient reports, including those in this study, are helpful in describing the long-term impacts of COVID-19 on pre-existing conditions given the changes to health management and individual behaviors during the pandemic, like seeking medical care for health complications. With the continued transmission of SARS-CoV-2 leading to new infections and reinfections, it will be important to continue assessing the long-term outcomes of those who experience exacerbated symptoms following their infection. New research questions and analyses may arise from the current analysis, including whether these pre-existing conditions are further exacerbated by reinfections or increase in frequency as new SARS-CoV-2 variants arise.

## Strengths and Limitations

The current analysis has several strengths. This report is one of the few of its kind to comprehensively describe the breadth of worsening conditions or concerns reported by U.S. adults after COVID-19 illness. We systematically categorized more than 100 chronic health conditions and concerns reportedly worsened after COVID-19 illness. Additionally, we incorporated narratives from respondents to contextualize and provide critical insight into how worsening conditions affect daily functioning and may translate into challenges in daily life. The use of self-report data in this study enabled information and qualitative details to be captured that are likely missing in electronic health records or other administrative data sources. These data provide context and information that has been missing from most COVID-19 literature.

This analysis also has some limitations. First, self-reported data may be subjected to recall or other bias. For example, respondents may not have reported a worsening condition because they did not remember or did not attribute the worsening of their condition or concerns to their COVID-19 illness. Alternatively, we cannot be sure all reported worsening conditions were attributed to COVID-19. Additionally, nearly 7% of the sample did not report a pre-existing health condition, making it difficult to account for their worsening condition. However, some respondents may not have reported a pre-existing health condition because it was not confirmed by a health professional. Lastly, the response rates differed by Track PCC site which may weaken the generalizability of these results.

## Conclusion

Although Long COVID research has primarily focused on incident conditions following COVID-19, our study examined reported exacerbation of various pre-existing conditions and concerns in the months following acute COVID-19 illness. These worsening conditions and concerns presented many reported challenges, including difficulties managing symptoms, having to address multiple worsening symptoms or conditions at once, and limitations in daily activities. Our findings suggest patients who have chronic conditions may need additional monitoring following COVID-19 to assess for exacerbation of pre-existing conditions.

## Data Availability

All data produced in the present study are available upon reasonable request to the authors.

## Declarations

This project was reviewed by each site’s Institutional Review Board and the Centers for Disease Control and Prevention and deemed a public health surveillance activity (PHSA). A PHSA does not meet the definition of human subject research and was conducted consistent with applicable federal law and policy. (§ See e.g., 45 C.F.R. part 46.102(f), 46.102(k), 46.102(1)(2), 21 C.F.R. part 56; 42 U.S.C. §241 (d); 5 U.S.C. §552a; 44 U.S.C. §3501 et seq.) Specifically, the Boise State University Institutional Review Board and the Temple University Institutional Review Board determined this project constitutes a PHSA. Subsequently, the University of Arizona Human Subjects Protection Program approved passive surveillance (#00001930) and active surveillance (#2003521636) as exempt research. The Indiana University Human Research Protection Program approved passive surveillance as exempt research (#16525) and active surveillance as expedited research (#20174). Active surveillance participants from all sites are given informed consent information before enrollment and consent is obtained from all adults and/or the parents/guardians of those□younger than 18 years and those□younger than 18 years provide assent per each site’s Institutional Review Board protocols.

The authors declare no competing interests.

## Appendix Complete list of worsening conditions after COVID-19 illness reported using free-text response by adult Track PCC respondents

1. **Respiratory**

- Respiratory/chest infections

`o` Pneumonia
`o` Bronchitis
`o` Influenza
- Respiratory signs/symptoms

`o` Difficulty breathing
`o` Shortness of breath
`o` Cough
`o` Runny/burning nose
`o` Sore throat
`o` Change in taste/smell
- Lung diseases and conditions

`o` Asthma
`o` Chronic obstructive pulmonary disease (COPD)
`o` Decreased lung function
`o` Respiratory failure
`o` Emphysema
`o` Lung scarring
- Chest concerns

`o` Chest inflammation/pain/burning sensation
`o` Chest heaviness
2. **General**

- Energy concerns

`o` Fatigue
`o` Tiredness
`o` Malaise
`o` No drive/low energy
`o` Lethargy
`o` Exertion
- Headache or migraine
- Chills
- Weight gain/loss
- Body weakness
- Hair loss
3. **Ears, Eyes, Nose and Throat**

- Eye concerns

`o` Orbital cellulitis
`o` Pink eye, conjunctivitis
`o` Poor eyesight, vision changes, blurry eyes
`o` Glaucoma
`o` Dry/sensitive eyes
- Ear concerns

`o` Ear infection
`o` Ringing ears/tinnitus
`o` Hearing loss
`o` Heat sensation behind ears
- Nose and Throat concerns

`o` Nosebleed
`o` Post-nasal drip
`o` Strep throat
`o` Voice changes
`o` Sinus condition/infection/headache
- Jaw/tooth concerns

`o` Tooth decay
`o` Temporomandibular joint (TMJ) disorder
4. **Musculoskeletal**

- Body aches/pain

`o` Joint aches/pain
`o` Arthritis
`o` Muscle aches/pain
`o` Soreness
- Inflammation/swelling
- Osteoporosis
5. **Cardiovascular/Cerebrovascular**

- Cardiac conditions

`o` Congestive heart failure (CHF)
`o` Atrial fibrillation (AFib)
`o` Heart attack
`o` Heart failure
`o` Heart disease
`o` Pulmonary hypertension
- Cardiac signs/symptoms
`o` Chest pounding
`o` Heart racing/palpitations
`o` Slowed heart rate
`o` Abnormal heart activity
- Vascular concerns

`o` Thoracic aortic aneurysm
`o` Deep Vein Thrombosis
`o` Poor circulation
- Cerebrovascular concerns

`o` Stroke
`o` Brain bleed
- Blood pressure changes
6. **Mental health**

- Anxiety/Depression

`o` Anxiety
`o` Depression
`o` Panic attacks
- Mental health signs/symptoms

`o` Stress
`o` Fearful
`o` Not feeling like myself
`o` Mood changes/swings
`o` Loss of interest
`o` Not enjoying life
`o` Poor emotional regulation
`o` Irritable
- Bipolar disorder
- Post-Traumatic Stress Disorder (PTSD)
7. **Neuropsychiatric**

- Sleep concerns

`o` Problems sleeping
`o` Insomnia
`o` Hypersomnolence
`o` Nightmares/bad dreams
- Memory issues

`o` Poor concentration
`o` Difficulty remembering
`o` Memory loss
`o` Mental clarity
`o` Brain fog
`o` Mental acuity
`o` Forgetfulness
`o` Mental decline
- Attention-Deficit/Hyperactivity Disorder (ADHD)
- Sleep apnea
- Dementia
8. **Immune/autoimmune**

- Illness frequency/duration changes

`o` Sick more often
`o` Longer recovery time
- Autoimmune disorders

`o` Immune thrombocytopenic purpura
`o` Lupus
`o` Celiac
`o` Sjogren’s
`o` Wegener’s
`o` Sclerosis
- Allergies
`o` Food allergies
`o` Seasonal allergies
- Spleen concerns
- Sarcoidosis
9. **Gastrointestinal (GI)**

- GI diagnoses

`o` Gastroesophageal Reflux Disease (GERD), gastritis, reflux
`o` Gastroparesis
`o` Esophagitis
`o` Intestinal bacteria
`o` Irritable Bowel Syndrome (IBS)
- GI signs/symptoms

`o` Stomach pain
`o` Diarrhea
`o` Constipation
- Change in appetite
- Pancreas concerns
- Gallstones
- Liver concerns
10. **Neurologic**

- Nerve concerns

`o` Nerve pain
`o` Nerve inflammation/swelling
`o` Neuropathy
- Brain conditions/disorders

`o` Traumatic Brain Injury (TBI)
`o` Seizures
`o` Arnold Chiari
- Dysautonomia, Postural orthostatic tachycardia syndrome (POTS)
- Balance difficulties

`o` Vertigo/dizziness
11. **Other Conditions**

- Hematologic cancers

`o` Leukemia
`o` Lymphoma
- Chronic Lyme disease
- Fibromyalgia
- Endometriosis
- Ehler’s Danlos
- Anemia
12. **Dermatologic/Skin**

- Skin diagnoses

`o` Psoriasis
`o` Shingles
`o` Eczema
- Hives
- Sores and rashes
13. **Urinary**

- Prostate concerns

`o` Prostate surgery
`o` Enlarged prostate
`o` Prostate cancer
- Kidney concerns
14. **Endocrine/Metabolic**

- Thyroid concerns

`o` Hypothyroidism
`o` Hashimoto’s
`o` Multinodular goiter
`o` Abnormal Thyroid-Stimulating Hormone (TSH) levels
- Hypokalemia
- Diabetes

