## Supplemental Table 1 for "Worsening pre-existing health conditions in U.S. adults with Long COVID"

**Supplementary Materials**

Table S1. Demographic characteristics of the overall Track PCC adult respondents

|  | **Adults enrolled in Track PCC by January 15, 2025** | **Adults reporting any worsening condition or concern** |
| --- | --- | --- |
| **N** | 2,227 | 263 |
| **Age, years (IQR)** | 57 (42, 68) | 57 (43, 67) |
| **Age group** |  |  |
| 18-49 years | 796 (35.7) | 89 (33.8) |
| 50-64 years | 680 (30.5) | 96 (36.5) |
| 65+ years | 751 (33.7) | 78 (29.7) |
| **Site** |  |  |
| Arizona | 762 (34.2) | 44 (16.7) |
| Comagine (The Bronx) | 156 (7.0) | 16 (6.1) |
| Indiana | 457 (20.5) | 70 (26.6) |
| Temple (Philadelphia) | 852 (38.3) | 133 (50.6) |
| **Sex** |  |  |
| Male | 627 (29.3) | 58 (22.2) |
| Female | 1,587 (71.7) | 203 (77.8) |
| *Missing* | *13* | *2* |
| **Race** |  |  |
| Black or African American | 480 (22.9) | 67 (26.7) |
| White or Caucasian | 1,343 (64.2) | 139 (55.4) |
| Any Other Race or Multiple Races | 269 (12.9) | 45 (17.9) |
| *Missing* | *135* | *13* |
| **Ethnicity** |  |  |
| Latinx/Hispanic | 361 (17.4) | 55 (22.4) |
| Non-Hispanic | 1,719 (82.6) | 191 (77.6) |
| *Missing* | *147* | *17* |
| **Pre-existing condition(s)** |  |  |
| Any underlying condition | 1,878 (85.1) | 246 (93.5) |
| *Missing* | 19 |  |
